# The estimated impact of mandatory front-of-pack nutrition labelling policies on adult obesity prevalence and cardiovascular mortality in England: a modelling study

**DOI:** 10.1101/2024.10.14.24315283

**Authors:** Rebecca Evans, Martin O’Flaherty, I Gusti Ngurah Edi Putra, Chris Kypridemos, Eric Robinson, Zoé Colombet

**Affiliations:** Department of Psychology, University of Liverpool, Liverpool, United Kingdom; Department of Public Health, Policy and Systems, University of Liverpool, Liverpool, United Kingdom

**Keywords:** microsimulation model, policy evaluation, inequalities, food labelling policies

## Abstract

**Objectives:** Since 2013, industry-endorsed front-of-pack traffic light labels have been implemented voluntarily on packaged food in the UK. The UK Government is now considering alternative labelling approaches which may be more effective, such as Chile’s mandatory nutrient warning labels. The primary aim of this study was to model the likely impact of implementing mandatory front-of-pack nutrition labels in England on energy intake and consequent population-level obesity, and, secondarily, cardiovascular disease (CVD) mortality.

**Design:** Microsimulation modelling analysis

**Setting:** England

**Model:** A microsimulation model (2024–2043) to estimate the impact of changing front-of-pack nutrition labels in England. The two main policy scenarios tested were mandatory implementation of (i) traffic light labels and (ii) nutrient warning labels. For each scenario, the impact of the policy through assumed changes in energy intake due to consumer behaviour change and reformulation was modelled.

**Main outcome measures:** Change in obesity prevalence (%) and CVD deaths prevented or postponed.

**Results:** Compared to the baseline scenario (current voluntary implementation of traffic light labelling), mandatory implementation of traffic light labelling was estimated to reduce obesity prevalence in England by 2.28% (95% UI –4.06 to –0.96) and prevent or postpone 17000 (95% UI 4700 to 48000) CVD deaths. Mandatory implementation of nutrient warning labelling was estimated to have a larger impact; a 3.68% (95% UI –9.94 to –0.18) reduction in obesity prevalence and the prevention/postponement of 29000 (95% UI 1200 to 110000) CVD deaths.

**Conclusions:** This work offers the first modelled estimation of the impact of introducing mandatory front-of-pack nutrition labels on health outcomes in the adult population in England. Findings suggest that mandatory implementation of nutrient warning labels would reduce rates of obesity and CVD deaths, compared to current voluntary or mandatory implementation of traffic light labelling, and should therefore be considered by the UK government.

**Funding:** European Research Council (Grant reference: PIDS, 8031940).

## Introduction

Diet-related disease is a major cause of poor population health and social inequalities in health (1). Many pre-prepared foods and non-alcoholic beverages (hereafter: food) are high in calories, added sugar, salt, and/or saturated fat (2,3). Excessive consumption of these nutrients increases the risk of obesity and other associated non-communicable diseases (NCD) such as cardiovascular disease (CVD), and NCD mortality (4).

In the UK, the average adult consumes an excess of 200-300 calories per day, and nearly two-thirds of UK adults are living with overweight or obesity (5,6). Notably, the prevalence of overweight and obesity is patterned by deprivation (14 percentage points higher in the most relative to the least deprived areas), and education (12 percentage points higher for those with no qualifications compared to those who are degree-level educated) (5). Therefore, there is a need for equitable public health policies that improve dietary quality across the population.

Front-of-pack nutrition labels are an evidence-based policy tool used to help consumers make healthier food choices and encourage industry to improve the nutritional profile of the products they sell (7). In the UK, an industry-endorsed traffic light front-of-pack nutrition label (see **Figure 1.A**) has been implemented voluntarily since 2013. This traffic light label uses green, amber, and red colours to indicate whether a product contains low, moderate, or high levels of nutrients of concern, alongside guideline daily amount (GDA) percentages for each nutrient (typically per serving). However, UK consumers report that the traffic light label is difficult to interpret, which may widen health inequalities (8). Additionally, less than half of consumers use the label to determine product calorie content, and calorie content specifically is not designated with a traffic light colour (9). It may be that simpler labels are required, as most consumers typically spend no more than a few seconds examining labels before making a food selection (10).

**Figure 1.**
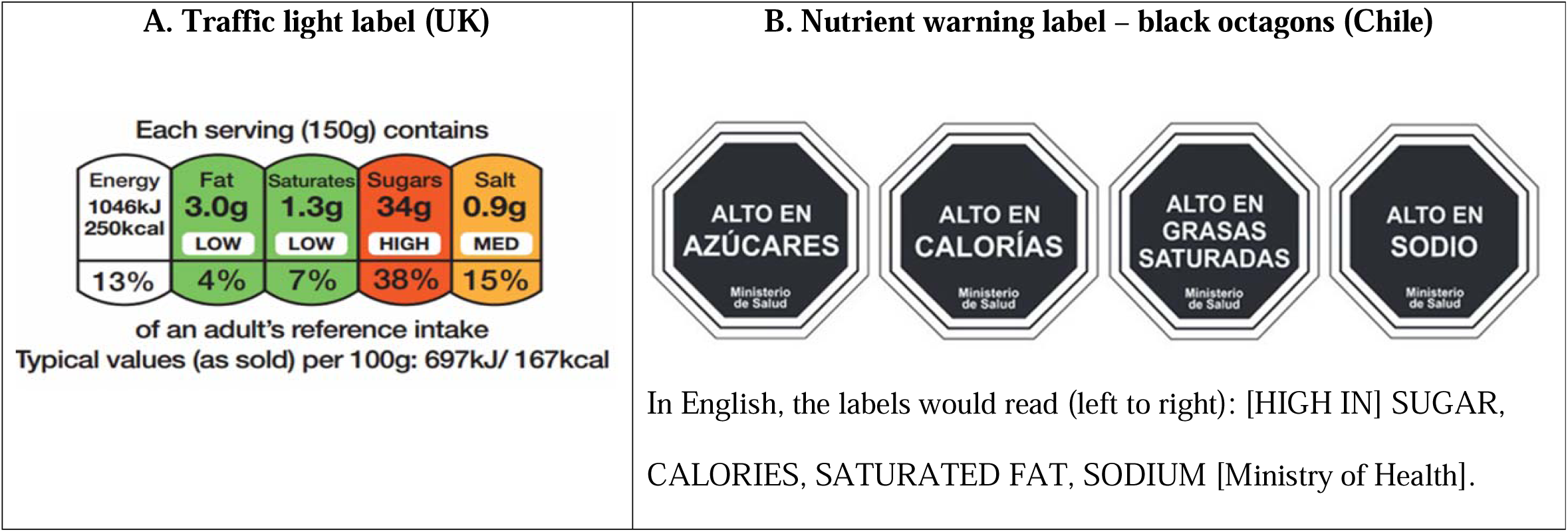
Front-of-pack nutrition labels examples.

In July 2020, the UK Government launched a consultation considering an alternative front-of-pack nutrition label to the traffic light (11). In the consultation, Chile’s nutrient warning labels were highlighted as a potential alternative, and the benefits of implementing mandatory front-of-pack labelling were discussed.

In 2016, Chile implemented a mandatory policy requiring packaged foods containing ‘high’ amounts (as defined by thresholds set by the Ministry of Health) of calories, added sugar, sodium, and/or saturated fat to display nutrient warning labels (12) (see **Figure 1.B**). Very similar policies have since been implemented in other South American countries, including Argentina, Brazil, Colombia, Mexico, Peru, and Uruguay (13,14). Mandatory nutrient warnings have also been implemented further afield in Canada and Israel, and policy development is under consideration in several other countries, including the US, India, and South Africa (15). Evidence indicates that implementation in Chile has reduced the purchase of energy (a relative 8.3% decrease, 95% CI: [5.0, 11.6]) and nutrients of concern (ranging from –9.6% for saturated fat to –20.2% for sugar) (16), and has led to product reformulation across all food groups, leading to reductions in energy content (–3.9%), and other labelled nutrients of concern (ranging from –1.5% for saturated fat to –15% for sugar) (17).

Furthermore, evidence from a meta-analysis of over 100 randomised controlled trials (RCTs) and quasi-experimental studies suggests that nutrient warning labels may perform better than traffic light labels in terms of reducing consumers’ purchase of energy (an additional 6.4% (95% CI: [0.4; 12.5] reduction) and nutrients of concern, and probability of choosing less healthy products (7). Therefore, it is important to examine the potential impact of their implementation in the UK on health outcomes such as adult obesity prevalence, to inform policy decision-making.

The present study aimed to estimate the likely long-term impacts of implementing (i) mandatory nutrient warning labels and (ii) mandatory traffic light labels on packaged in-store foods, relative to the current voluntary implementation of traffic light labels, on energy intake and consequent population-level obesity prevalence and cardiovascular mortality due to change in BMI in England.

## Methods

### Model overview

We built a dynamic, discrete-time, stochastic, open-cohort microsimulation model to quantify the estimated effects of implementing front-of-pack nutrition labels in England; an adaptation of the IMPACT NCD Model based on the IMPACT Food Policy Model (18). The model simulates the life-course of individuals and their counterfactuals under alternative policy scenarios. This enables the detailed simulation of diet policies and their impact on relevant exposures, subsequent disease epidemiology, and mortality in a competing risk framework that accounts for different lag-times between exposures and outcomes. In this case, we simulated the effects of implementing mandatory front-of-pack nutrition labels (nutrient warning and traffic light) on daily energy intake from packaged food, and subsequent population-level obesity prevalence and CVD mortality due to change in BMI. We modelled the population of England, aged 30 to 89 years, over 20 years (2024 to 2043) using a synthetic population stratified by age, sex and Index of Multiple Deprivation (IMD) that captures the real demographics, energy intakes, and disease epidemiology of the actual population of England using available national data sources (see below and in Appendix section “Creation of our synthetic population”).

We evaluated two main policy scenarios:

1. Traffic light labels are implemented as a mandatory policy
2. Nutrient warning labels are implemented as a mandatory policy

We compared each scenario with a counterfactual “no intervention” (baseline) scenario, which corresponds to the current England legislation: continued voluntary implementation of traffic light labels.

We did not model the impact of Nutri-Score, an alternative front-of-pack label which uses a colour spectrum and letter grades to summarise product healthiness, as a main scenario (19), This is because meta-analytic evidence suggests that it does not perform significantly differently to the traffic light label in terms of reducing energy purchased (7). Instead, results for Nutri-Score are presented in the Appendix (see **Appendix Table 4**).

### Front-of-pack nutrition labels

Front-of-pack nutrition labels impact diet through (1) consumer behaviour change, and (2) industry response, i.e., reformulation of the products by industry (see **Figure 2**).

**Figure 2.**
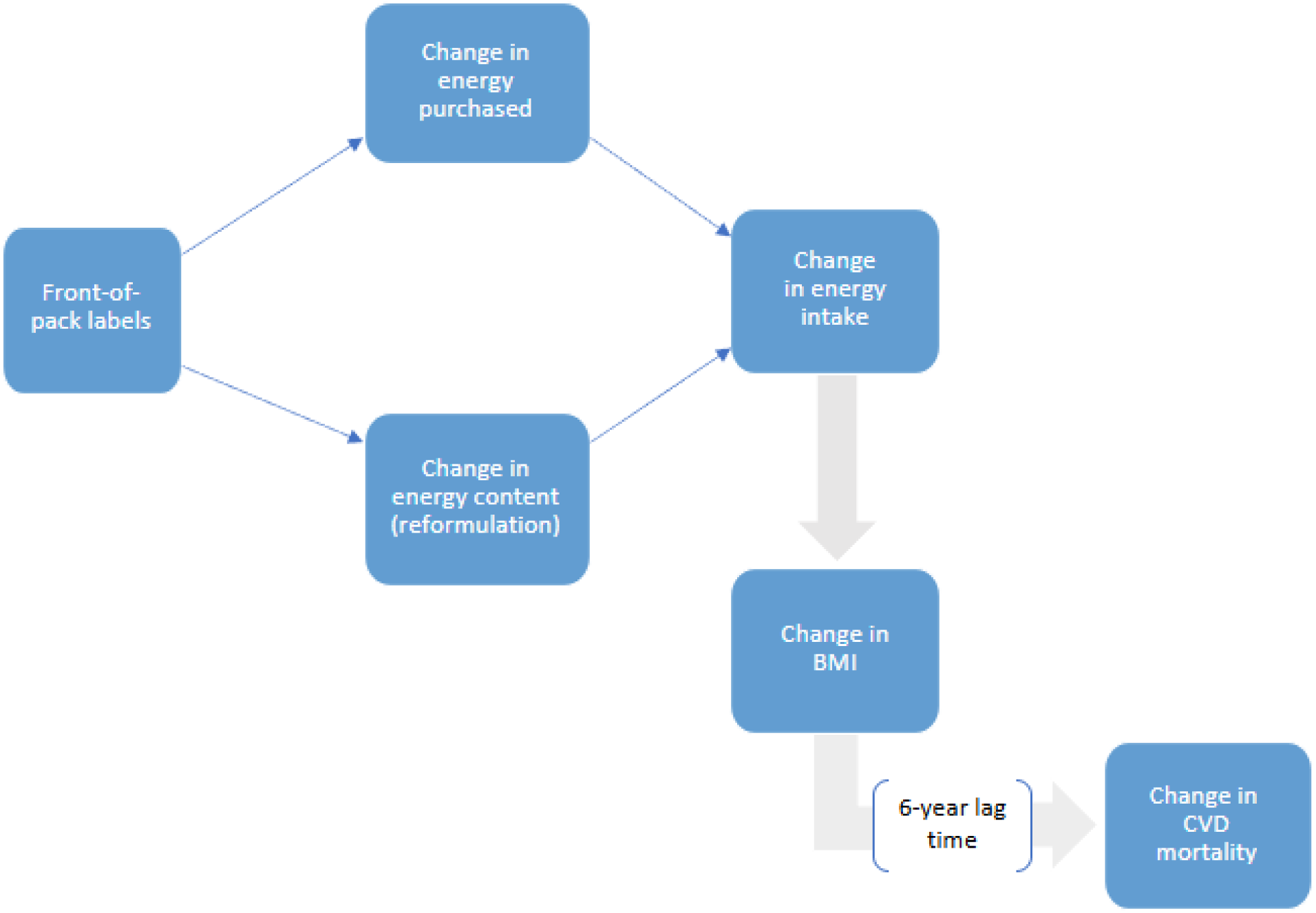
Logic diagram of the impact of front-of-pack labelling on obesity prevalence and cardiovascular disease (CVD) mortality. Abbreviation: BMI: Body Mass Index

### Effect on consumer behaviour change

We assumed that the traffic light labels and nutrient warning labels would reduce energy purchased from packaged food by 6.5% (95% CI: [2.0; 11.0]), 12.9% (95% CI: [8.0; 18.0]), and 6% (95% CI: [1.0; 11.0]) respectively, compared to no label, based on the estimates from Song et al.’s review and network meta-analysis (7). Based on the same meta-analysis, we assume that nutrient warning labels will outperform traffic light labels in reducing the total amount of energy purchased by 6.4% (95% CI: [0.4; 12.5]). Based on existing literature, we assumed no differential policy effects by sex, age or socioeconomic position (7,20). Due to an absence of evidence, we assumed both labels have a consistent effect on consumer behaviour over time.

### Effect on energy content reformulation

For nutrient warning labels, we assumed a 3.9% (95% CI: [12.5; 4.95]) reduction in energy content of labelled packaged foods, based on evidence from Chile post-implementation (17). While there is no available data specifically in relation to traffic light labelling and product reformulation, evidence suggests that a small amount of reformulation does occur in response to food labelling, particularly when it is implemented mandatorily (21–23). Therefore, we also assumed the same 3.9% reduction in energy content of packaged foods in response to mandatory traffic light labelling.

### Label coverage

We assumed that all packaged products (100%) would feature a traffic light label, as under mandatory implementation, this would be required by law (16). Under current voluntary implementation, it is estimated that 75% of packaged products feature the label (24), so mandatory implementation would yield an additional 25% coverage. For nutrient warning labels, based on evidence on the proportion of products featuring a “high in” warning in Chile, we assumed that 51% (95% CI: [49.0; 52.0]) of packaged foods in England would feature the label (i.e., will be above threshold for warning) (25). The nutritional quality of packaged food in Chile is relatively similar to the UK; the average Health Star Rating for packaged food is 2.44 compared to 2.83 (scores range from 0.5 to 5, with a higher score indicating better nutritional quality) (26). Moreover, an analysis of food items from the UK NDNS indicated that approximately 40% of UK food items meet requirements for a red traffic light label, and this figure does not include items that would be labelled due to being high in energy (27). Research suggests that 32% of UK supermarket snack foods alone exceed adult energy intake recommendations (3) and therefore it is reasonable to estimate that this would amount to at least an additional 10% of products being labelled, consistent with the 51% figure derived from Chile.

### Estimating model uncertainty

We used the Monte Carlo approach (100 iterations) to estimate the uncertainty of model parameters. The sources of uncertainty we considered were the uncertainty of the relative risk of coronary heart disease (CHD) and stroke based on BMI, the uncertainty of mortality forecasts, and the uncertainty of the policy (label) effect. We summarised the output distributions by reporting the medians and 95% uncertainty intervals (UIs).

### One-way sensitivity analyses on key parameters

#### Change in nutrient warning labels coverage

Evidence from Chile suggests that approximately one year after initial implementation of the nutrient warning label policy, reformulation resulted in a decrease in the proportion of products featuring a label from 51% to 44% (95% CI: [42.0 – 45.0]) (25). Reformulation to reduce nutrients of concern is consistently observed in response to the introduction of front-of-pack nutrition labelling policies in various countries, including Australia, Canada, the Netherlands, and New Zealand, to avoid a “negative” label (e.g., a low health rating) or the absence of a “positive” label (e.g., a healthy choice indicator) (28). Therefore, in this sensitivity analysis we assume that coverage is 51% for the first-year post-implementation, and coverage then drops to 44% thereafter.

#### Chile’s black octagon specifically (as opposed to nutrient warning labels more generally)

In this sensitivity analysis, we test based on evidence from Chile specifically, post-implementation (as opposed to meta-analytic data on nutrient warning labels in general from experimental studies), which suggests an overall 8.8% (95% CI: [-7.1 to –10.5]) reduction in energy purchased (16). Notably, nutrient warning labels were introduced in Chile as part of a set of policies, including restrictions on food marketing to children, and therefore this reduction in energy purchase may not me wholly attributable to nutrient warning label implementation.

#### Lower reformulation due to traffic light labels

It is possible that reformulation of energy content may be lower in response to traffic light labelling relative to nutrient warning labelling. This is because calories are not colour-coded in traffic light labels and therefore food companies may be less inclined to reformulate energy content of products. We assumed there would be a smaller 0.9% (95% CI [-3.1, 4.9]) reduction in energy content, based on a meta-analysis of food labelling effects on product energy reformulation (23).

A further detailed description of the model, input sources, and key assumptions are provided in the **Appendix**.

### Model engine

Front-of-pack nutrition labels are hypothesised to reduce energy intake, which will subsequently impact the body weight of the population (i.e., BMI), and, in turn, change CVD mortality risk. This pathway is described in **Figure 2** and detail in **Appendix** (section “Estimating the effect of change in energy intake upon obesity prevalence and CVD mortality”). In short, the change in energy intake is calculated by subtracting intake post-intervention from baseline intake for each year. Changes in energy intake are then converted into changes in body weight, based on principles of energy conservation, using the Christiansen & Garby prediction formula (29) (detail in **Appendix** section “Estimating the effect of change in energy intake on BMI”). The estimated change in BMI is then calculated based on the estimated change in body weight, which allows us to estimate the change in obesity prevalence. Next, these changes in BMI are used to estimate changes in CVD mortality risk, with a 6-year lag time (30) (see details in **Appendix** section “Estimating the effect of change in BMI upon CVD mortality”). Using this information, new mortality rates and, consequently, the number of deaths projected can be estimated.

### Model outputs

The model produced the change in obesity prevalence and the total number of deaths prevented or postponed (DPPs) for each scenario. The equity impact of the intervention was examined by calculating the ratio between the most and least deprived quintile groups (using the IMD). Results are presented for English adults aged 30 to 89 years from 2024 to 2043, rounded to 2 significant figures for mortality and rounded to 2 decimal places for obesity prevalence.

### Data sources

We constructed a synthetic population of England to simulate the population-level impact of the policy scenarios. This is described in the Appendix section “Data sources used in our model” and **Appendix Table 1**. The England population projections were derived from the Office for National Statistics (ONS), and mortality trend projections were based on the CVD deaths observed in England from 1981 to 2016.

**Table 1:** Summary of key model assumptions.

|  | Traffic light label | Nutrient warning label |
| --- | --- | --- |
| *Main assumptions* |  |  |
| Effect on energy intake | -6.5% [-11%; -2%] (7) | -12.9% [-18%; -8%]<br>(outperforms the traffic light label by 6.4% [0.4; 12.5] (7) |
| Effect on reformulation in terms of energy content | -3.9% [-12.5; 4.95] (17) | -3.9% [-12.5; 4.95] (17) |
| Label coverage on packaged products | 100% (currently 75% under voluntary implementation) (24) | 51% [49%; 52%] (25) |
| *Sensitivity assumptions* |  |  |
| Changes in label coverage over time due to reformulation | - | Drops to 44% [42.0; 45.0] 4 years post-implementation (25) |
| Chile's black octagon nutrient warning label effectiveness on energy intake from labelled products | - | -8.8% [7.1.; -10.5] (16) |
| Effect on reformulation in terms of energy content | -0.9% [-3.1, 4.9] (23) | - |

We used generalised additive models for location, shape and scale (GAMLSS) to estimate (i) BMI and (ii) energy intake distributions dependent on age, sex, and IMD. GAMLSS can handle complex relationships between the response variable and its predictors and numerous types of distributions (31). Trends in energy intake daily energy intakes and BMI were obtained from the nationally representative National Diet and Nutrition Survey (NDNS) 2009-2019. These trends in energy intake and BMI observed in the last 10 years in England were assumed to continue in the future. To obtain the daily energy from packaged food bought from grocery retail stores, we assumed that 55% of all food and beverage expenditure (including alcoholic beverages) was for at-home consumption (vs. 45% spent on restaurants and other out-of-home food services) (32) and that 80% of the products purchased are packaged (vs. 20% fresh) (8) (see details in **Appendix** section “Modelling approach and scenarios”).

R (version 4.3.0) was used to conduct all data management and statistical analyses. We used the “demography” package (33) for forecasting mortality and the “gamlss” package to fit the distribution (34). For code, see https://github.com/zoecolombet/FoPLabels_code

## Results

Maintaining current voluntary traffic light labelling would result in obesity prevalence of 28.03% (95% UI 27.74 – 28.30) by 2043.

The implementation of mandatory traffic light labelling in England was estimated to reduce obesity prevalence by 1.49 percentage points (absolute; 95% UI –2.44 to –0.76; **Table 2**) in the next 20 years when only considering consumer behaviour change (i.e., change in energy intake). Reformulation of the energy content of the packaged products sold was estimated to lower obesity prevalence by 0.66 percentage points (95% UI –2.79 to 0.00; **Table 2**). Combining these factors would result in a decrease of 2.28 percentage points in obesity prevalence among adults (95% UI –4.06 to –0.96; **Table 2**).

**Table 2:**
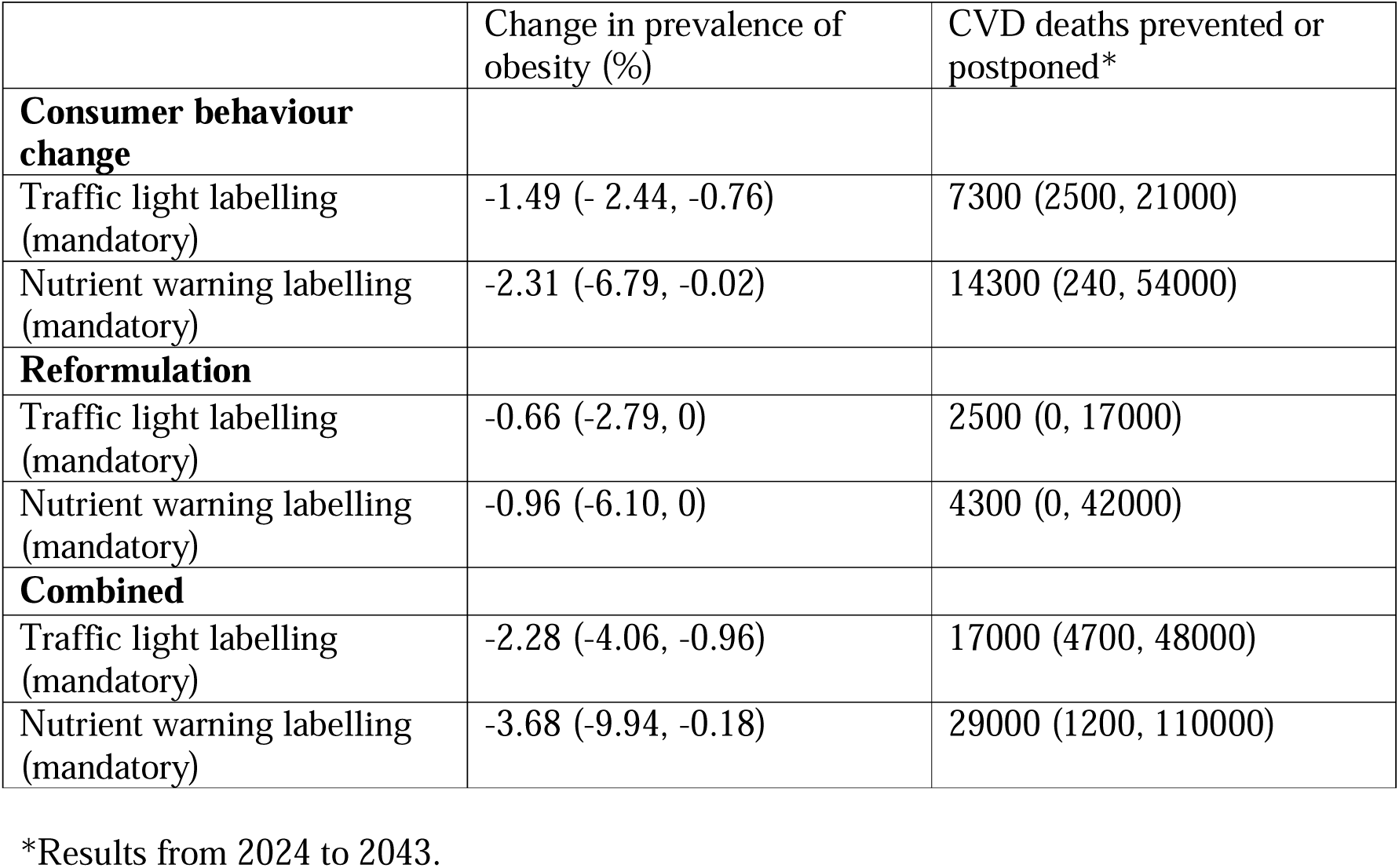
Estimated change in obesity prevalence and CVD mortality due to change in BMI in adults in England (2024–43), according to different front-of-pack labelling implementation scenarios.

Implementing mandatory nutrient warning labels on packaged products was estimated to have a larger impact and reduce obesity prevalence by 2.31 percentage points (95% UI –6.79 to – 0.02; **Table 2**) when only considering consumer behaviour change. Reformulation of the energy content of the packaged products sold was estimated to lower obesity prevalence by 0.96 percentage points (95% UI –6.10 to 0; **Table 2**). Combining these factors would result in a decrease of 3.68 percentage points in obesity prevalence among adults (95% UI –9.94 to – 0.18; **Table 2**).

Maintaining current voluntary implementation of traffic light labelling in England, the current cardiovascular mortality trends were estimated to result in approximately 1,900,000 deaths (95% UI 1,100,000 – 3,300,000) in English adults by 2043.

Implementing traffic light labelling mandatorily would prevent or postpone approximately 7300 deaths (95% UI 2500 to 21000; **Table 2**) attributable to BMI-related CVD, based on consumer behaviour change alone. Reformulation was estimated to avert 2500 deaths (95% UI 0 to 17000; **Table 2**). Combined, this would result in 17000 deaths (95% UI 4700 to 48000; **Table 2**) prevented or postponed.

Again, implementing mandatory nutrient warning labels was estimated to have a larger impact, resulting in the prevention or postponement of an estimated 14300 (95% UI 240 to 54000) deaths based on consumer behaviour change, 4300 deaths (95% UI 0 to 42000; **Table 2**) based on reformulation, and 29000 deaths (95% UI 1200 to 110000; **Table 2**) based on the two combined.

The introduction of either front-of-package label as a mandatory policy is estimated to reduce obesity prevalence and relative CVD deaths to a similar extent across socioeconomic deprivation levels (see **Table 3**).

**Table 3:** Estimated change in obesity prevalence and CVD mortality due to change in BMI in adults in England (2024–43), according to IMD quintile groups and different front-of-pack labelling implementation scenarios.

|  | Prevalence of obesity,<br>percentage points | CVD deaths |
| --- | --- | --- |
|  | Predicted obesity<br>prevalence | CVD deaths predicted |
| **Current voluntary traffic light**<br>**labelling** |  |  |
| *Q1 (most deprived)* | 32.53 (32.00, 33.04) | 470,000 (270,000 –<br>830,000) |
| *Q5 (least deprived)* | 24.29 (23.55, 24.85) | 290,000 (170,000 – 500,<br>000) |
|  | Predicted change in<br>obesity prevalence | CVD deaths prevented or<br>postponed |
| **Mandatory traffic light labelling –**<br>**consumer behaviour change** |  |  |
| *Q1* | -1.46 (-2.24, -0.71) | 2000 (240, 5500) |
| *Q5* | -1.48 (-2.46, -0.75) | 1000 (0, 4500) |
| **Mandatory traffic light labelling -**<br>**reformulation** |  |  |
| *Q1* | -0.66 (-2.85, 0) | 500 (0, 6500) |
| *Q5* | -0.65 (-2.73, 0) | 250 (0, 2000) |
| **Mandatory traffic light labelling -**<br>**combined** |  |  |
| *Q1* | -2.14 (-3.96, -0.91) | 4000 (740, 14000) |
| *Q5* | -2.28 (-4.08, -0.93) | 2500 (500, 8000) |
| **Mandatory nutrient warning**<br>**labelling – consumer behaviour**<br>**change** |  |  |
| *Q1* | -2.25 (-6.25, -0.01) | 3500 (0, 13000) |
| *Q5* | -2.31 (-6.81, -0.03) | 2000 (0, 8800) |
| **Mandatory nutrient warning**<br>**labelling - reformulation** |  |  |
| *Q1* | -0.90 (-5.58, 0) | 1000 (0, 12000) |
| *Q5* | -1.05 (-6.20, 0) | 500 (0, 5500) |
| **Mandatory nutrient warning**<br>**labelling - combined** |  |  |
| *Q1* | -3.61 (-9.58, -0.19) | 7500 (0, 30000) |
| *Q5* | -3.59 (-9.80, -0.20) | 4500 (0, 18000) |

See **Appendix Table 3** for sensitivity analysis results relating to nutrient warning label coverage, Chile’s nutrient warning label specifically, and traffic light label reformulation. Briefly, nutrient warning labels with reduced coverage, and Chile’s warning label specifically still outperformed traffic light labels. Traffic light labels saw a notable decrease in performance using the more conservative reformulation estimate. See **Appendix Table 4** for results relating to Nutri Score. As expected, results for Nutri Score were very similar to those for traffic light labelling.

## Discussion

This work offers the first modelled estimation of the impact of changing front-of-pack nutrition label policy on obesity prevalence and CVD mortality in the adult population in England. Our findings indicate that, in place of current voluntary traffic light labelling, the introduction of mandatory nutrient warning labels would reduce obesity prevalence and CVD deaths substantially more than making traffic light labels mandatory, with no differential effects on health inequalities.

Our findings are largely consistent with the existing limited evidence in this area. One previous study modelled the impact of nutrient warning labels in Mexico (35). The study estimated a mean caloric reduction of 36.8 kcal/day/person, and, 5 years post-implementation, 1.3 million fewer cases of obesity (5% reduction). A handful of studies have modelled the impact of traffic light labelling on NCD mortality. One study modelling impact in Canada (36) estimated that 11715 deaths per year due to diet-related NCDs, and 10490 deaths per year due to energy intake alone would be prevented. However, this was contingent on Canadians using the traffic light labelling to avoid foods labelled with red lights. Another study estimated the impact of Nutri-Couleurs (traffic light label) across 27 EU nations and found no significant effect on NCD mortality (37). However, the effect estimate for change in energy intake was derived from a large-scale randomised controlled trial in French supermarkets which only covered four product types (bread, ready meals, fresh catering, and pastries) (38), as opposed to the use of meta-analytic evidence in the present research.

Although the current research provides important insights into the likely impact of changing front-of-pack nutrition label policy in England, there are limitations to be acknowledged. We assumed that reductions in energy intake would be in response to labelled products, which may be an overestimate for traffic light labels as not all products would feature a “red” indicator. We also assumed that energy intake trends from NDNS will continue, but it is possible that COVID-19 and/or the cost-of-living crisis may result in long-term changes. Our results will also underestimate total policy benefits as we did not include changes in childhood obesity in our model.

It is also important to acknowledge that the present research underestimates the impact of the labelling policies on total CVD mortality as due to model design we do not model effects of policies due to changes in intake of nutrients of concern (salt, sugar, saturated fat) and instead model change via energy intake and reductions to BMI. Excess intake of salt, sugar, and saturated fat is associated with CVD risk (39). Evidence suggests that labelling policies decrease the purchase of nutrients of concern, especially nutrient warning labels relative to traffic light labels, so impacts on CVD mortality are likely to be particularly underestimated for nutrient warning labels (7,20).

We did not model a scenario where nutrient warning labels are implemented voluntarily, as there are no examples of such implementation. Moreover, the current evidence suggest that voluntary, industry-endorsed initiatives in the context of front-of-package labelling are likely to be ineffective for several reasons, such as industry manipulation of label design, noncompliance (particularly as nutrient warning labels are known to deter purchase of labelled products), and a lack of independent target setting, monitoring, and enforcement (40,41)Finally, while nutrient warning labels appear effective in reducing purchase and intake of energy and nutrients of concern, it may be that alternative/additional labels are required to encourage consumers to select health protective food options (i.e., those that contain nutrients that the population do not consume enough of, e.g., fiber, vitamin D).

Several assumptions in our model were constrained by a lack of available evidence and these areas might benefit from further research. Firstly, there was no available data on how the effect of the label on consumer behaviour change may change over time. Theoretically, if people become habituated to front of pack labels, then the effect may decrease, or conversely, if nutrient literacy and awareness strengthen over time then the effect may increase (8). Secondly, there was no available data on compensatory effects from intake of fresh food in place of packaged food, or intake from out-of-home eating. Thirdly, although there is some self-report evidence to suggest that age, education, and ethnicity may impact understanding of, and therefore response to traffic light labels (8), there was no consistent evidence that demographic factors moderate the effect of labels on product choice (7,20).

The World Health Organization (WHO) does not at present recommend the use of any specific labelling scheme but encourages research institutions and member states to continue analysing information to inform decisions (42). This new modelled evidence supports the use of nutrient warning labels to reduce population-level obesity. While such labels are gaining global popularity, the UK and Europe are yet to adopt this policy approach. It is recommended that the UK Government replaces its current voluntary traffic light labelling system with mandatory nutrient warning labelling to reduce rates of obesity and related CVD deaths.

## Conclusion

Mandatory implementation of nutrient warning labels appears to be the most favorable policy option for the UK government to substantially reduce rates of obesity, compared to current voluntary or mandatory implementation of traffic light labelling.

## Declarations

### Data sharing

ONS and NDNS data are available online. The “demography” package for R software has been used for forecasting mortality and the “gamlss” package has been used to fit the distribution. Syntax for the generation of derived variables and for the analysis used in this study are available publicly: https://github.com/zoecolombet/FoPLabels_code

## Funding

Salaries for ZC and ER were fully and part-funded, respectively, by the European Research Council under the European Union’s Horizon 2020 research and innovation programme (Grant reference: PIDS, 803194). ER and RE are funded by the National Institute for Health and Care Research (NIHR) Oxford Health Biomedical Research Centre (BRC) (Grant reference: NIHR203316).

## Role of the funding source

The funder played no role in the study design, data collection, data analysis, data interpretation, writing of the paper, or the decision to submit this work for publication.

## Competing interest statement

All authors have completed the Unified Competing Interest Form and declare: no support from any organisation for the submitted work; no financial relationships with any organisations that might have an interest in the submitted work in the previous three years; no other relationships or activities that could appear to have influenced the submitted work.

## Transparency declaration

The lead author (R.E) affirms that the manuscript is an honest, accurate, and transparent account of the study being reported; that no important aspects of the study have been omitted; and that any discrepancies from the study as planned have been explained.

## Copyright statement

The Corresponding Author (R.E.) has the right to grant on behalf of all authors and does grant on behalf of all authors, an exclusive licence on a worldwide basis to the BMJ Publishing Group Ltd to permit this article to be published in BMJ editions and any other BMJPGL products and sublicences such use and exploit all subsidiary rights, as set out in our licence.

## Ethical approval

Ethical approval was not required for this study.

## Authors’ contributions

ZC, RE, ER, MO’F, and EP designed the study. ZC and RE directly accessed and verified the underlying data reported in this article. ZC and RE developed the model. CK, MO’F, and ER supervised ZC and RE. RE and ZC did the analysis and drafted the manuscript. All authors contributed to the data interpretation and revised each draft for important intellectual content. All authors had final responsibility for the decision to submit for publication.

## Data Availability

ONS and NDNS data are available online. The demography package for R software has been used for forecasting mortality and the gamlss package has been used to fit the distribution. Syntax for the generation of derived variables and for the analysis used in this study are available publicly: https://github.com/zoecolombet/FoPLabels_code

## Abbreviations

BMI: Body mass index
CVD: Cardiovascular disease
IMD: Index of Multiple Deprivation
NCD: Non-communicable diseases

## Notes

### Competing Interest Statement

The authors have declared no competing interest.

## References

1. Gilmore AB, Fabbri A, Baum F, Bertscher A, Bondy K, Chang HJ, et al. Defining and conceptualising the commercial determinants of health. The Lancet. 2023 Apr;401(10383):1194– 213.

2. Huang Y, Burgoine T, Essman M, Theis DRZ, Bishop TRP, Adams J. Monitoring the Nutrient Composition of Food Prepared Out-of-Home in the United Kingdom: Database Development and Case Study. JMIR Public Health Surveill. 2022 Sep 8;8(9):e39033.

3. Marty L, Evans R, Sheen F, Humphreys G, Jones A, Boyland E, et al. The energy and nutritional content of snacks sold at supermarkets and coffee shops in the UK. J Human Nutrition Diet. 2021 Dec;34(6):1035–41.

4. Anand SS, Hawkes C, De Souza RJ, Mente A, Dehghan M, Nugent R, et al. Food Consumption and its Impact on Cardiovascular Disease: Importance of Solutions Focused on the Globalized Food System. Journal of the American College of Cardiology. 2015 Oct;66(14):1590–614.

5. Baker C. Research Briefing – Obesity statistics [Internet]. House of Commons Librar; 2023 [cited 2024 Apr 17]. Available from: https://researchbriefings.files.parliament.uk/documents/SN03336/SN03336.pdf

6. Tedstone A, Targett V, Mackinlay B, Owtram G, Coulton V, Morgan K, et al. Calorie reduction: The scope and ambition for action [Internet]. Public Health England; 2018 [cited 2024 Apr 17]. Available from: https://assets.publishing.service.gov.uk/government/uploads/system/uploads/attachment_data/file/800675/Calories_Evidence_Document.pdf

7. Song J, Brown MK, Tan M, MacGregor GA, Webster J, Campbell NRC, et al. Impact of color-coded and warning nutrition labelling schemes: A systematic review and network meta-analysis. Ares G, editor. PLoS Med. 2021 Oct 5;18(10):e1003765.

8. Osman M, Jenkins S. Consumer responses to food labelling: A rapid evidence review [Internet]. Food Standards Agency; 2021 [cited 2024 Apr 17]. Available from: https://www.food.gov.uk/research/consumer-responses-to-food-labelling-a-rapid-evidence-review

9. Food Standards Agency. Eating Well Choosing Better Tracker Survey Wave 8 2022 [Internet]. 2022. Available from: https://www.food.gov.uk/research/ewcb-2022-results

10. Sanjari SS, Jahn S, Boztug Y. Dual-process theory and consumer response to front-of-package nutrition label formats. Nutrition Reviews. 2017 Nov 1;75(11):871–82.

11. GOV.UK. Front-of-pack nutrition labelling in the UK: building on success [Internet]. 2020. Available from: https://www.gov.uk/government/consultations/front-of-pack-nutrition-labelling-in-the-uk-building-on-success

12. Reyes M, Garmendia ML, Olivares S, Aqueveque C, Zacarías I, Corvalán C. Development of the Chilean front-of-package food warning label. BMC Public Health. 2019 Dec;19(1):906.

13. Taillie LS, Hall MG, Gómez LF, Higgins I, Bercholz M, Murukutla N, et al. Designing an Effective Front-of-Package Warning Label for Food and Drinks High in Added Sugar, Sodium, or Saturated Fat in Colombia: An Online Experiment. Nutrients. 2020 Oct 13;12(10):3124.

14. White M, Barquera S. Mexico Adopts Food Warning Labels, Why Now? Health Systems & Reform. 2020 Dec;6(1):e1752063.

15. UNC Global Food Research Program. Front-of-Package (FOP) Food Labelling: Empowering consumers and promoting healthy diets [Internet]. Available from: https://www.globalfoodresearchprogram.org/wp-content/uploads/2022/10/FOP_Factsheet_HSR_update.pdf

16. Taillie LS, Bercholz M, Popkin B, Rebolledo N, Reyes M, Corvalán MC. Decreases in purchases of energy, sodium, sugar, and saturated fat 3 years after implementation of the Chilean food labeling and marketing law: An interruptd time series analysis. PLOS Medicine. 2024 Sep 27;21(9):e1004463.

17. Quintiliano Scarpelli D, Pinheiro Fernandes AC, Rodriguez Osiac L, Pizarro Quevedo T. Changes in Nutrient Declaration after the Food Labeling and Advertising Law in Chile: A Longitudinal Approach. Nutrients. 2020 Aug 8;12(8):2371.

18. Pearson-Stuttard J, Bandosz P, Rehm CD, Penalvo J, Whitsel L, Gaziano T, et al. Reducing US cardiovascular disease burden and disparities through national and targeted dietary policies: A modelling study. PLoS Med. 2017 Jun;14(6):e1002311.

19. Santé publique France. NUTRI-SCORE Questions & Answers English version [Internet]. 2024. Available from: https://www.santepubliquefrance.fr/content/download/150263/file/QR_scientifique_technique_EN_12052020.pdf

20. Croker H, Packer J, Russell SJ, Stansfield C, Viner RM. Front of pack nutritional labelling schemes: a systematic review and meta-analysis of recent evidence relating to objectively measured consumption and purchasing. J Human Nutrition Diet. 2020 Aug;33(4):518–37.

21. Ganderats-Fuentes M, Morgan S. Front-of-Package Nutrition Labeling and Its Impact on Food Industry Practices: A Systematic Review of the Evidence. Nutrients. 2023 Jun 5;15(11):2630.

22. Vandevijvere S, Vanderlee L. Effect of Formulation, Labelling, and Taxation Policies on the Nutritional Quality of the Food Supply. Curr Nutr Rep. 2019 Sep;8(3):240–9.

23. Shangguan S, Afshin A, Shulkin M, Ma W, Marsden D, Smith J, et al. A Meta-Analysis of Food Labeling Effects on Consumer Diet Behaviors and Industry Practices. American Journal of Preventive Medicine. 2019 Feb;56(2):300–14.

24. House of Commons Health Committee. Childhood obesity— brave and bold action [Internet]. 2015. Available from: https://publications.parliament.uk/pa/cm201516/cmselect/cmhealth/465/465.pdf

25. Reyes M, Smith Taillie L, Popkin B, Kanter R, Vandevijvere S, Corvalán C. Changes in the amount of nutrient of packaged foods and beverages after the initial implementation of the Chilean Law of Food Labelling and Advertising: A nonexperimental prospective study. Wareham NJ, editor. PLoS Med. 2020 Jul 28;17(7):e1003220.

26. Dunford EK, Ni Mhurchu C, Huang L, Vandevijvere S, Swinburn B, Pravst I, et al. A comparison of the healthiness of packaged foods and beverages from 12 countries using the Health Star Rating nutrient profiling system, 2013–2018. Obesity Reviews. 2019 Nov;20(S2):107–15.

27. Dicken SJ, Batterham RL, Brown A. Nutrients or processing? An analysis of food and drink items from the UK National Diet and Nutrition Survey based on nutrient content, the NOVA classification and front of package traffic light labelling. Br J Nutr. 2024 May 14;131(9):1619–32.

28. Roberto CA, Ng SW, Ganderats-Fuentes M, Hammond D, Barquera S, Jauregui A, et al. The Influence of Front-of-Package Nutrition Labeling on Consumer Behavior and Product Reformulation. Annu Rev Nutr. 2021 Oct 11;41(1):529–50.

29. Christiansen E, Garby L. Prediction of body weight changes caused by changes in energy balance. Eur J Clin Invest. 2002 Nov;32(11):826–30.

30. Emerging Risk Factors Collaboration, Wormser D, Kaptoge S, Di Angelantonio E, Wood AM, Pennells L, et al. Separate and combined associations of body-mass index and abdominal adiposity with cardiovascular disease: collaborative analysis of 58 prospective studies. Lancet. 2011 Mar 26;377(9771):1085–95.

31. Stasinopoulos MD, Rigby RA, Heller GZ, Voudouris V, Bastiani FD. Flexible Regression and Smoothing: Using GAMLSS in R. New York: Chapman and Hall/CRC; 2017. 571 p.

32. Statista. Share of household food and drink expenditure in the United Kingdom (UK) from 2000 to 1st quarter 2020, by at-home and out-of-home consumption. Available from: https://www.statista.com/statistics/941699/in-home-versus-out-of-home-food-and-drink-spending-united-kingdom-uk/

33. Hyndman R, Booth H, Tickle L, Maindonald J. Package ‘demography’ for R [Internet]. [cited 2022 Nov 9]. Available from: https://github.com/robjhyndman/demography

34. Stasinopoulos M, Rigby R, Voudouris V, Akantziliotou C, Enea M, Kiose D, et al. Package ‘gamlss’: Generalized Additive Models for Location Scale and Shape [Internet]. 2024 [cited 2024 Apr 24]. Available from: https://cran.r-project.org/web/packages/gamlss/gamlss.pdf

35. Basto-Abreu A, Torres-Alvarez R, Reyes-Sánchez F, González-Morales R, Canto-Osorio F, Colchero MA, et al. Predicting obesity reduction after implementing warning labels in Mexico: A modeling study. Clément K, editor. PLoS Med. 2020 Jul 28;17(7):e1003221.

36. Labonté ME, Emrich TE, Scarborough P, Rayner M, L’Abbé MR. Traffic light labelling could prevent mortality from noncommunicable diseases in Canada: A scenario modelling study. Vadiveloo MK, editor. PLoS ONE. 2019 Dec 27;14(12):e0226975.

37. Devaux M, Aldea A, Lerouge A, Vuik S, Cecchini M. Establishing an EU-wide front-of-pack nutrition label: Review of options and model-based evaluation. Obesity Reviews. 2024 Jun;25(6):e13719.

38. Allais O, Albuquerque P, Bonnet C, Dubois P. Évaluation Expérimentation Logos Nutritionnels Rapport pour le FFAS [Internet]. 2017. Available from: https://sante.gouv.fr/IMG/pdf/rapport_final_groupe_traitement_evaluation_logos.pdf

39. Bowen KJ, Sullivan VK, Kris-Etherton PM, Petersen KS. Nutrition and Cardiovascular Disease—an Update. Curr Atheroscler Rep. 2018 Jan 30;20(2):8.

40. Laverty AA, Kypridemos C, Seferidi P, Vamos EP, Pearson-Stuttard J, Collins B, et al. Quantifying the impact of the Public Health Responsibility Deal on salt intake, cardiovascular disease and gastric cancer burdens: interrupted time series and microsimulation study. J Epidemiol Community Health. 2019 Sep;73(9):881–7.

41. Knai C, Petticrew M, Douglas N, Durand MA, Eastmure E, Nolte E, et al. The Public Health Responsibility Deal: Using a Systems-Level Analysis to Understand the Lack of Impact on Alcohol, Food, Physical Activity, and Workplace Health Sub-Systems. IJERPH. 2018 Dec 17;15(12):2895.

42. World Health Organisation. Guiding principles and framework manual for front-of-pack labelling for promoting healthy diets [Internet]. 2019 [cited 2024 Jul 15]. Available from: https://www.who.int/publications/m/item/guidingprinciples-labelling-promoting-healthydiet

