## Appendix for "The estimated impact of mandatory front-of-pack nutrition labelling policies on adult obesity prevalence and cardiovascular mortality in England: a modelling study"

### **Front-of-pack nutrition labels**

### ***Modelling approach and scenarios***

We built a dynamic, discrete-time, stochastic, open-cohort microsimulation model to quantify the potential population-level impacts of the implementation of different front-of-pack nutrition labels in England; an adaptation of the IMPACT NCD Model based on the IMPACT Food Policy Model (1). The model simulates the life-course of individuals and their counterfactuals under alternative policy scenarios. This enables the detailed simulation of diet policies and their impact on relevant exposures, subsequent disease epidemiology, and mortality in a competing risk framework that accounts for different lag-times between exposures and outcomes. We quantified the impact of mandatory implementation of (i) traffic light labels and (ii) nutrient warning labels, relative to a “no intervention” baseline scenario which corresponds to the current voluntary implementation of traffic light labels in the UK. We estimated the impact of this policy over a 20-year horizon from 2022 to 2043 in the adult population of England, aged 30-89.

*Assumptions*

Coverage: We assumed that 55% of all food and beverage expenditure (including alcoholic beverages) was for at-home consumption (vs. 45% spent on restaurants and other out-of-home food services) (2), and that 80% of the products purchased are packaged (vs. 20% fresh) (3). We assumed that traffic light labels would feature on 100% of packaged products due to mandatory implementation; current estimates indicate that traffic light labels feature on 75% of packaged products, so this amounts to an additional 25% coverage (4). We assumed that nutrient warning labels would feature on 51% (95% CI: [49.0; 52.0]) of packaged products, based on evidence from Chile (5). The nutritional quality of packaged food in Chile is relatively similar to the UK; the average Health Star Rating for packaged food is 2.44 compared to 2.83 (scores range from 0.5 to 5, with a higher score indicating better nutritional quality (6). Moreover, an analysis of food items from the UK NDNS indicated that approximately 40% of UK food items meet requirements for a red traffic light label, but this figure does not include items that would be labelled due to being “high in” energy (7). Research suggests that 32% of UK supermarket snack foods alone exceed adult energy intake recommendations (8) and therefore it is reasonable to estimate that this could amount to at least an additional 10% of packaged food being labelled, consistent with the 51% figure derived from Chile.

Effects on intake: From the literature, front-of-pack nutrition labels seem to impact energy intake through (i) consumer behaviour change (i.e., customers opt for healthier or lower-calorie options) and (ii) industry response (i.e., food reformulation to reduce calorie or nutrient of concern content).

We assumed that the traffic light labels, and nutrient warning labels would reduce total energy purchased by 6.5% (95% CI: [2.0; 11.0]), and 12.9% (95% CI: [8.0; 18.0]) respectively, compared to no label, based on the estimates from Song et al.’s review and network meta-analysis (9). Based on the same meta-analysis, we assume that the nutrient warning labels will outperform traffic light labels in reducing the total amount of energy purchased by 6.4% (95% CI: [0.4; 12.5]). We assumed that a decrease in energy purchased equates to a decrease in energy consumed, and therefore traffic light labelling would decrease total energy consumed by 6.5% (95% CI: [2.0; 11.0]), and nutrient warning labels by a further 6.4% (95% CI: [0.4; 12.5]) on top of this. Based on the literature, we assumed no differential policy effects by sex, age or socioeconomic position (9,10). Due to an absence of evidence, we assume both labels have a consistent effect on consumer behaviour over time. We opted to model data based on Song et al. (9) rather than Croker et al. (10) as this meta-analysis is more up to date, more comprehensive, and reports relative change in energy intake (% change) compared to change in energy intake per 100g.

While to date there are no meta-analyses examining the impact of Chile’s black octagon labels specifically, there is emerging evidence of its effects post-implementation. Therefore, we also examine the potential impact of implementing the black octagon labels specifically (as opposed to nutrient warning labels more generally). We assume an 8.8% (95% CI: [-7. 1 to –10.5]) reduction in energy intake based on purchasing evidence post-implementation in Chile (11).

Evidence suggests that reformulation does occur in response to food labelling, particularly when it is implemented mandatorily (12–14). We assume a 3.9% (95% CI: [12.5; 4.95]) reduction in energy content based on evidence post-implementation of nutrient warning labels in Chile (15) . We make the same assumption for both nutrient warning labels and traffic light labels, as data for traffic light labelling specifically is lacking.

Evidence suggests that because of product reformulation in response to implementation of nutrient warning labels in Chile, over time fewer packaged products meet the threshold for featuring a warning label. Specifically, evidence from Chile suggests that after initial implementation of the nutrient warning label policy, reformulation resulted in a decrease in the proportion of products featuring a label, to 44% (95% CI: [42.0 - 45.0]) (5). Therefore, for nutrient warning labels, we assumed that coverage is 51% for the first 4 years post-implementation, and following reformulation, this drops to 44%.

In our model, we compared two main scenarios to the baseline scenario, and conducted several sensitivity analyses:

Scenario 1: Traffic light labelling is implemented as a mandatory policy

Consumer behaviour change: We assumed that traffic light labels would reduce total energy purchased by 6.5% (95% CI: [2.0; 11.0] based on Song et al.’s meta-analysis (9). We assumed that traffic light labels would feature on 100% of packaged products due to mandatory implementation; current estimates indicate that traffic light labels feature on 75% of packaged products, so this amounts to an additional 25% coverage (4). Therefore, the -6.5% energy purchase effect is applied to 25% of packaged products.

Reformulation: There is no traffic light label–specific figure for reformulation of energy content, but evidence suggests that some reformulation does occur in response to mandatory front-of-package food labelling (12). Therefore, we assumed a 3.9% (95% CI: [12.5; 4.95]) reduction in energy content of the 25% additional packaged products now featuring a traffic light label, based on evidence from nutrient warning labelling post-implementation in Chile (15).

Consumer behaviour change + reformulation: We combined the consumer behaviour change and reformulation assumptions described above.

Sensitivity analyses

Reformulation: It is possible that reformulation of energy content may be lower in response to traffic light labelling relative to nutrient warning labelling. This is because calories are not colour-coded in traffic light labels (i.e., even if “high” this wouldn’t be highlighted in red), and therefore food companies may be less inclined to reformulate their products. Based on this, we assumed a lower 0.9% (95% CI [-3.1, 4.9]) reduction in energy content of the 25% additional packaged products now featuring a traffic light label, based on a meta-analysis of general food labelling effects on reformulation (13).

Scenario 2: Nutrient warning labelling is implemented as a mandatory policy

Consumer behaviour change: We assumed that nutrient warning labels would reduce total energy purchased by 12.9% (95% CI: [8.0; 18.0]), based on Song et al’s meta-analysis (9). Based on the same meta-analysis, we assumed that the nutrient warning labels would outperform traffic light labels in reducing the total amount of energy purchased by 6.4% (95% CI: [0.4; 12.5]). We assumed that nutrient warning labels would feature on 51% (95% CI: [49.0; 52.0]) of packaged products, based on evidence from Chile (5). Therefore, the -6.4% energy purchase effect is applied to 51% of packaged products.

Reformulation: We assumed a 3.9% (95% CI: [12.5; 4.95]) reduction in energy content of the 51% of packaged foods that would feature a nutrient warning label, based on evidence from Chile post-implementation (15).

Consumer behaviour change + reformulation: We combined the consumer behaviour change and reformulation assumptions described above.

Sensitivity analyses:

Coverage: Evidence from Chile suggests that after initial implementation of the nutrient warning label policy, reformulation resulted in a decrease in the proportion of products featuring a label, to 44% (95% CI: [42.0 - 45.0]) one-year post-implementation (5). Therefore, we assumed that coverage is 51% for the first-year post-implementation, and following reformulation, this drops to 44%.

Chile’s black octagon: We assume an 8.8% (95% CI: [-7.1 to –10.5]) reduction in energy intake based on evidence from Chile specifically (11).

Nutri-Score:

Consumer behaviour change: We assumed that Nutri-Score labels would reduce total energy purchased by 6% (95% CI: [1.0; 11.0]), compared to no label, based on the estimates from Song et al.’s review and network meta-analysis (9). Based on the same meta-analysis, we assume that the Nutri-Score labels will not outperform traffic light labels in reducing the total amount of energy purchased (only a nonsignificant 1% further reduction (95% CI: [0.05; -0.04]). We assumed that Nutri-Score labels would feature on 100% of packaged products due to mandatory implementation; current estimates indicate that traffic light labels feature on 75% of packaged products, so this amounts to an additional 25% coverage (4). Therefore, the -6% energy purchase effect is applied to 25% of packaged products.

Reformulation: We assumed a 3.9% (95% CI: [12.5; 4.95]) reduction in energy content of the 25% additional packaged products now featuring a Nutri-Score label, based on evidence from nutrient warning labelling post-implementation in Chile (15).

Consumer behaviour change + reformulation: We combined the consumer behaviour change and reformulation assumptions described above.

### **Creation of our synthetic population**

We constructed a synthetic population of England to simulate the population-level impact of the policy scenarios. The data sources used in our model are presented in **Appendix Table 1**, and the key assumptions we made in this simulation modelling are displayed in **Appendix Table 2**.

#### ***Population projection***

The Office for National Statistics (ONS) (16,17) provided the population and population projections for England to 2043, and this was stratified by sex and age. The ONS does not provide population estimates and projections by Index of Multiple Deprivation (IMD). Therefore, we assumed that the relative difference in population estimates across IMD quintiles by age and sex between 2022 and 2043 was equal to the relative difference in 2020. The English population composition by age, sex, and IMD was imported from ONS (18).

#### ***CVD Mortality projection***

We projected mortality trends to 2043, by age, sex, and IMD quintiles, based on the number of annual CVD observed by ONS for England from 1981 to 2016. This number consisted of CHD (ICD-10: I20 to I25) and overall strokes (ICD-10: I60 to I69, I64, I69.4, and I69.8) (19). Projecting mortality based on previous trends allowed us to consider declining CVD mortality trends, meaning that we could avoid overestimating the benefits of any CVD intervention (1).

#### ***Energy intake from purchased packaged products***

We estimated the energy intake from packaged products purchased using the representative National Diet and Nutrition Survey (NDNS) 2009-2019 (Years 1 to 11) (20). Data indicates that 55% of all food and beverage expenditure is for at-home consumption (vs. Out-of-home food services) (2), and 80% of purchased food products are packaged (vs. fresh) (3). Therefore, we used the formula (daily energy intake [NDNS] *0.55 *0.80) to determine energy intake from purchased packaged products.

We used generalised additive models for location, shape and scale (GAMLSS) to estimate energy intake distributions dependent on age, sex, and IMD. GAMLSS can handle complex relationships between the response variable and its predictors and numerous types of distributions (23).

#### ***BMI***

We also used GAMLSS to estimate BMI distributions dependent on age, sex, and IMD. Trends in BMI were obtained from the nationally representative National Diet and Nutrition Survey (NDNS) 2009-2019. We assumed that the trends in BMI observed in the last 10 years in England will continue in the future.

### **Estimating the effect of change in energy intake on CVD mortality**

#### ***Estimating the change in energy intake***

Implementing front-of-pack nutrition labels will impact energy purchase and then energy intake. To calculate population-level change in energy intake due to the implementation of these labels, we subtracted energy intake post-policy from baseline intake for each year, assuming that all energy purchased will be eaten. We assumed that the implementation of the policy would immediately impact energy intake, and this effect would remain consistent throughout the simulation period, as we did not have further evidence to the contrary.

#### ***Estimating the effect of change in energy intake on BMI***

Changes in energy intake will subsequently impact BMI. We used a prediction formula by Christiansen & Garby (developed based on principles of energy conservation) to convert the change in energy intake into a change in body weight. The formula is as follows:

$$\Delta BW = k * \Delta(\frac{Energy intake}{Physical activity level})$$

Body weight (Δ BW) is in kilograms (kg); Energy intake is in MegaJoule (MJ); Physical activity level (PAL) is calculated as the total energy expenditure divided by the resting energy expenditure; and 𝑘 is a constant value based on (i) fundamental principles of energy conservation and (ii) directly measured data (constant values of 17.7 and 20.7 for men and women, respectively) (21).

PAL was kept constant at 1.5 (21) to represent limited physical activity (22) under the assumption that implementing front-of-package labels has no impacts on physical activity levels. The estimated change in body weight informed the potential change in BMI under the assumption that individuals’ height remained unchanged.

***Estimating the effect of change in BMI upon CVD mortality***

The Emerging Risk Factors Collaboration (ERFC) (23) computed the risks of CVD for one standard deviation (SD) increase in BMI (4.56 kg/m²). The risks were estimated for individuals with a BMI ≥ 20 kg/m² in that study, so we calculated the baseline and new risks for CVD for each one SD increase in BMI. We assigned a risk of 1 to individuals with BMI < 20 kg/m². The risks for the remaining individuals were decided based on age-specific estimates from the ERFC, which are adjusted for sex and smoking status (23), in addition to the average BMI.

We computed the population-attributable risk fraction (PARF), which is the proportion of disease-specific mortality (i.e., CVD mortality) that is attributable to a specific risk factor (i.e., BMI ≥ 20 kg/m²).

Firstly, in a microsimulation context in which the risk factor is known at an individual level, PARF can be estimated using the formula:

$$PARF=1- \frac{n}{\sum_{i=1}^{n} {RR}_{BMI, i}}$$

*n* represents the number of synthetic individuals in the population, and *RR_BMI_* refers to the relative risk of the risk factor (BMI ≥ 20 kg/m²) associated with CVD mortality for each individual (*i*).

Secondly, the proportion of CVD mortality that is not attributable to modelled risk factor(s) can be estimated using the formula:

$$M_{Theoretical minimun}= M_{Observed}\times(1-PARF)$$

$M_{Observed}$ is the observed CVD mortality, and *PARF* is from the previous calculation. $M_{Theoretical minimum}$ represents the CVD mortality if all modelled risk factors were at optimal levels. This is calculated by age, sex, and IMD for all years of the simulation, but it is assumed that *PARF* remains stable after the initial year.

Thirdly, if we assume that $M_{Theoretical minimum}$ is the annual baseline probability of a synthetic individual to die of CVD for a given age, sex, IMD, and due to risk factors not in the model (e.g., genetics, smoking etc.), then we can estimate the individualised annual probability of CVD mortality given their risk factors using the formula:

$$P\left( CVD | age, sex, IMD, nonmodelled exposures, BMI \right)= M_{Theoretical minimum}\times{RR}_{BMI, i}$$

As previously mentioned, ${RR}_{BMI}$ refers to the relative risk related to the synthetic individual's specific BMI exposure.

We presented the above for simplicity using CVD as one disease. In reality we do these calculations separately for CHD and ischaemic stroke deaths and we present their sum as CVD deaths.

By subtracting the number of CVD deaths in a policy scenario (that assumes a counterfactual energy intake) from the baseline scenario we calculate the number of deaths prevented or postponed (DPPs). We then aggregate the DPPs over the simulation period (20 years from 2024 to 2043) and present the cumulative number of DPPs.

We assumed that a change in energy intake would affect the change in BMI in less than a year. This was not assumed for the impact of change in BMI on CVD mortality risk. Instead, we implemented a 6-year lag time for the change in BMI to affect CVD mortality risk. This is consistent with the median duration (5.7 years) reported by ERFC for developing the first CVD-related outcome (14). Based on this, in our model, the BMI change from 2024 was only modelled to impact CVD mortality risk from 2030. This means that the reported DPPs cover a 14 –year-period toward the end of the modelling horizon.

### **Estimating model uncertainty**

We used the 2^nd^ order Monte Carlo approach with 100 iterations (29) to estimate the uncertainty of model parameters. There are different possible sources of uncertainty; uncertainty of the relative risk of (i) coronary heart disease (CHD) and (ii) stroke mortality based on BMI, uncertainty of mortality forecasts, and uncertainty of the front-of-pack label effect. Results were presented as the median alongside 95% uncertainty intervals (UIs).

### **Appendix Table 1.** Data sources used in the model.

| **Parameter** | **Outcome** | **Details** | **Differences by sociodemographic groups** | **Source** | **Projection distribution of the mean** | **Uncertainty** |
| --- | --- | --- | --- | --- | --- | --- |
| Population size estimates | Population | ONS data from 2001 to 2020 | Stratified by region, year, age, sex | ONS population estimates for England (16) | - | - |
| Population composition | Population | IMD repartition in the England population in 2020 | Stratified by age, sex, IMD | ONS for England (18) | - | - |
| Population projection | Population | 2020-2120 England population projection produced by the ONS | Stratified by year, age, sex | ONS population projection for England (17) | - | No uncertainty assumed for population forecast |
| Mortality | Deaths from CVD | Underlying cause of death: England 1981-2016 | Stratified by year, age, sex, IMD, and cause of death | ONS cause of death for England 1981-2016 (19) | Log normal | Mean ± SD |
| Mean energy intake from packaged food products | Mean energy intake | NDNS 2009-2019 | Stratified by year, age, sex, and IMD | NDNS 2009-2019 (20) | Log normal | Mean ± SD |
| Effect of traffic light label | Change in energy purchase/ intake | An estimate from a meta-analysis of over 100 randomised controlled trials (RCTs) and quasi-experimental studies | No differential effect | Song et al. (9) | - | Mean & 95% CI |
| Effect of nutrient warning label | Change in energy purchase/intake | An estimate from a meta-analysis of over 100 randomised controlled trials (RCTs) and quasi-experimental studies | No differential effect | Song et al. (9) | - | Mean & 95% CI |
| Effect of the policy on product reformulation | Change in energy purchase/ intake | A longitudinal analysis of food nutritional labelling declarations from 70% of the most consumed packaged foods in Chile | No differential effect | Scarpelli et al. (15) | - | Mean & 95% CI |
| Effect of change in energy intake on BMI | Change in BMI |  | Stratified by sex | Christiansen & Garby (21) | - | - |
| Effect of change in BMI on CVD mortality risk | Change in CVD mortality | A collaborative analysis of 58 prospective studies | Stratified by age | Emerging Risk Factors Collaboration (23), Figure 2 | Log normal | Mean ± SD |

BMI: body mass index; CVD: cardiovascular diseases; IMD: Index of Multiple Deprivation; NDNS: National Diet and Nutrition Survey; ONS: Office for National Statistics; SD: standard deviation.

### **Appendix Table 2.** Assumptions implemented in the model.

Key assumptions

- The composition of the population IMD in 2020 will be the same from 2024 – 2043

- There will be no change in the effect of the labels over time

- There will be no difference in the effect of labels according to age, sex, and IMD

- Energy purchase is equivalent to energy intake

- Compliance with a mandatory front-of-pack labelling policy will be 100%

- 80% of food products in retail stores are packaged

- 55% of all food expenditure is from retail stores for at-home consumption

**Appendix Table 3**: Sensitivity analyses of the estimated change in obesity prevalence and CVD mortality due to change in BMI in adults in England (2024–43), according to different front-of-pack labelling implementation scenarios

|  | Change in prevalence of obesity, percentage points | CVD deaths prevented or postponed* |
| --- | --- | --- |
| *Nutrient warning labels* |  |  |
| Coverage | -2.31 (- 6.82, 0) | 12000 (0, 46000) |
| Chile’s label specifically (consumer behaviour change) | -3.30 (-8.00, -0.44) | 16000 (2000, 60000) |
| *Traffic light labels* |  |  |
| Reformulation | -0.19 (-1.08, 0) | 500 (0, 7500) |

### **Appendix Table 4**: Estimated change in obesity prevalence and CVD mortality due to change in BMI in adults in England (2024–43), according to implementation of the Nutri-Score

|  | Change in prevalence of obesity, percentage points | CVD deaths prevented or postponed* |
| --- | --- | --- |
| *Nutri-Score* |  |  |
| Consumer behaviour change | -1.43 (-2.35, -0.31) | 7000 (1500, 20000) |
| Reformulation | -0.79 (-2.64, 0) | 4000 (0, 16000) |
| Combined | -2.13 (-4.26, -0.74) | 18000 (4000, 41000) |

**References**

1.	Pearson-Stuttard J, Bandosz P, Rehm CD, Penalvo J, Whitsel L, Gaziano T, et al. Reducing US cardiovascular disease burden and disparities through national and targeted dietary policies: A modelling study. PLoS Med. 2017 Jun;14(6):e1002311.

2.	Statista. Share of household food and drink expenditure in the United Kingdom (UK) from 2000 to 1st quarter 2020, by at-home and out-of-home consumption. Available from: https://www.statista.com/statistics/941699/in-home-versus-out-of-home-food-and-drink-spending-united-kingdom-uk/

3.	Osman M, Jenkins S. Consumer responses to food labelling: A rapid evidence review [Internet]. Food Standards Agency; 2021 [cited 2024 Apr 17]. Available from: https://www.food.gov.uk/research/consumer-responses-to-food-labelling-a-rapid-evidence-review

4.	House of Commons Health Committee. Childhood obesity— brave and bold action [Internet]. 2015. Available from: https://publications.parliament.uk/pa/cm201516/cmselect/cmhealth/465/465.pdf

5.	Reyes M, Smith Taillie L, Popkin B, Kanter R, Vandevijvere S, Corvalán C. Changes in the amount of nutrient of packaged foods and beverages after the initial implementation of the Chilean Law of Food Labelling and Advertising: A nonexperimental prospective study. Wareham NJ, editor. PLoS Med. 2020 Jul 28;17(7):e1003220.

6.	Dunford EK, Ni Mhurchu C, Huang L, Vandevijvere S, Swinburn B, Pravst I, et al. A comparison of the healthiness of packaged foods and beverages from 12 countries using the Health Star Rating nutrient profiling system, 2013–2018. Obesity Reviews. 2019 Nov;20(S2):107–15.

7.	Dicken SJ, Batterham RL, Brown A. Nutrients or processing? An analysis of food and drink items from the UK National Diet and Nutrition Survey based on nutrient content, the NOVA classification and front of package traffic light labelling. Br J Nutr. 2024 May 14;131(9):1619–32.

8.	Marty L, Evans R, Sheen F, Humphreys G, Jones A, Boyland E, et al. The energy and nutritional content of snacks sold at supermarkets and coffee shops in the UK. J Human Nutrition Diet. 2021 Dec;34(6):1035–41.

9.	Song J, Brown MK, Tan M, MacGregor GA, Webster J, Campbell NRC, et al. Impact of color-coded and warning nutrition labelling schemes: A systematic review and network meta-analysis. Ares G, editor. PLoS Med. 2021 Oct 5;18(10):e1003765.

10.	Croker H, Packer J, Russell SJ, Stansfield C, Viner RM. Front of pack nutritional labelling schemes: a systematic review and meta‐analysis of recent evidence relating to objectively measured consumption and purchasing. J Human Nutrition Diet. 2020 Aug;33(4):518–37.

11.	Taillie LS, Bercholz M, Popkin B, Rebolledo N, Reyes M, Corvalán MC. Decreases in purchases of energy, sodium, sugar, and saturated fat 3 years after implementation of the Chilean food labeling and marketing law: An interruptd time series analysis. PLOS Medicine. 2024 Sep 27;21(9):e1004463.

12.	Ganderats-Fuentes M, Morgan S. Front-of-Package Nutrition Labeling and Its Impact on Food Industry Practices: A Systematic Review of the Evidence. Nutrients. 2023 Jun 5;15(11):2630.

13.	Shangguan S, Afshin A, Shulkin M, Ma W, Marsden D, Smith J, et al. A Meta-Analysis of Food Labeling Effects on Consumer Diet Behaviors and Industry Practices. American Journal of Preventive Medicine. 2019 Feb;56(2):300–14.

14.	Vandevijvere S, Vanderlee L. Effect of Formulation, Labelling, and Taxation Policies on the Nutritional Quality of the Food Supply. Curr Nutr Rep. 2019 Sep;8(3):240–9.

15.	Quintiliano Scarpelli D, Pinheiro Fernandes AC, Rodriguez Osiac L, Pizarro Quevedo T. Changes in Nutrient Declaration after the Food Labeling and Advertising Law in Chile: A Longitudinal Approach. Nutrients. 2020 Aug 8;12(8):2371.

16.	Office for National Statistics (ONS). Estimates of the population for the UK, England and Wales, Scotland and Northern Ireland [Internet]. 2021 [cited 2022 Jun 15]. Available from: https://www.ons.gov.uk/peoplepopulationandcommunity/populationandmigration/populationestimates/datasets/populationestimatesforukenglandandwalesscotlandandnorthernireland

17.	Office for National Statistics (ONS). Principal projection - England summary [Internet]. 2022 [cited 2022 Jun 15]. Available from: https://www.ons.gov.uk/peoplepopulationandcommunity/populationandmigration/populationprojections/datasets/tablea14principalprojectionenglandsummary

18.	Office for National Statistics (ONS). Populations by Index of Multiple Deprivation (IMD) decile, England and Wales, 2020 [Internet]. 2021 [cited 2022 Jun 15]. Available from: https://www.ons.gov.uk/peoplepopulationandcommunity/populationandmigration/populationestimates/adhocs/13773populationsbyindexofmultipledeprivationimddecileenglandandwales2020

19.	Office for National Statistics (ONS). Deaths by selected causes and populations, both by deprivation decile areas, 5 year age groups and sex, England and Wales, registered years 1981 to 2016 [Internet]. 2018 [cited 2022 Nov 8]. Available from: https://www.ons.gov.uk/peoplepopulationandcommunity/birthsdeathsandmarriages/deaths/adhocs/008024deathsbyselectedcausesandpopulationsbothbydeprivationdecileareas5yearagegroupsandsexenglandandwalesregisteredyears1981to2016

20.	University of Cambridge, NatCen Social Research. National Diet and Nutrition Survey. UK Data Service; 2019.

21.	Christiansen E, Garby L. Prediction of body weight changes caused by changes in energy balance. Eur J Clin Invest. 2002 Nov;32(11):826–30.

22.	World Health Organization (WHO), Food and Agriculture Organization of the United Nations, United Nations University. Human Energy Requirements: Report of a Joint FAO/WHO/UNU Expert Consultation. Rome: Food & Agriculture Org.; 2004. 96 p.

23.	Emerging Risk Factors Collaboration, Wormser D, Kaptoge S, Di Angelantonio E, Wood AM, Pennells L, et al. Separate and combined associations of body-mass index and abdominal adiposity with cardiovascular disease: collaborative analysis of 58 prospective studies. Lancet. 2011 Mar 26;377(9771):1085–95.

24.	Khosravi A, Nazemipour M, Shinozaki T, Mansournia MA. Population attributable fraction in textbooks: Time to revise. Global Epidemiology. 2021 Nov 1;3:100062.

25.	Crockett RA, King SE, Marteau TM, Prevost AT, Bignardi G, Roberts NW, et al. Nutritional labelling for healthier food or non-alcoholic drink purchasing and consumption. Cochrane Database Syst Rev. 2018 Feb 27;2:CD009315.

26.	Shangguan S, Afshin A, Shulkin M, Ma W, Marsden D, Smith J, et al. A Meta-Analysis of Food Labeling Effects on Consumer Diet Behaviors and Industry Practices. Am J Prev Med. 2019 Feb;56(2):300–14.

27.	Petimar J, Zhang F, Rimm EB, Simon D, Cleveland LP, Gortmaker SL, et al. Changes in the calorie and nutrient content of purchased fast food meals after calorie menu labeling: A natural experiment. PLOS Medicine. 2021 Jul 12;18(7):e1003714.

28.	Zlatevska N, Neumann N, Dubelaar C. Mandatory Calorie Disclosure: A Comprehensive Analysis of Its Effect on Consumers and Retailers. Journal of Retailing. 2018 Mar 1;94(1):89–101.

29.	Koerkamp BG, Stijnen T, Weinstein MC, Hunink MGM. The Combined Analysis of Uncertainty and Patient Heterogeneity in Medical Decision Models. Med Decis Making. 2011 Jul 1;31(4):650–61.
